# SARS-CoV-2 patient self-testing with an antigen-detecting rapid test: a head-to-head comparison with professional testing

**DOI:** 10.1101/2021.01.06.20249009

**Authors:** Andreas K. Lindner, Olga Nikolai, Chiara Rohardt, Franka Kausch, Mia Wintel, Maximilian Gertler, Susen Burock, Merle Hörig, Julian Bernhard, Frank Tobian, Mary Gaeddert, Federica Lainati, Victor M. Corman, Terry C. Jones, Jilian A. Sacks, Joachim Seybold, Claudia M. Denkinger, Frank P. Mockenhaupt

## Abstract

**Background:** Antigen-detecting rapid diagnostic tests (Ag-RDTs) have been widely recommended as a complement to RT-PCR. Considering the possibility of nasal self-sampling and the ease-of-use in performing the test, self-testing may be an option.

**Methods and Findings:** We performed a manufacturer-independent, prospective diagnostic accuracy study of nasal mid-turbinate self-sampling and self-testing when using a WHO-listed SARS-CoV-2 Ag-RDT. Symptomatic participants suspected to have COVID-19 received written and illustrated instructions. Procedures were observed without intervention. For comparison, Ag-RDTs with nasopharyngeal sampling were professionally performed. Estimates of agreement, sensitivity, and specificity relative to RT-PCR on a combined oro-/nasopharyngeal sample were calculated. Feasibility was evaluated by observer and participant questionnaires.

Among 146 symptomatic adults, 40 (27.4%) were RT-PCR-positive for SARS-CoV-2. Sensitivity with self-testing was 82.5% (33/40 RT-PCR positives detected; 95% CI 68.1-91.3), and 85.0% (34/40; 95% CI 70.9-92.9) with professional testing. The positive percent agreement between self-testing and professional testing on Ag-RDT was 91.4% (95% CI 77.6-97.0), and negative percent agreement 99.1% (95% CI 95.0-100). At high viral load (>7.0 log_10_ SARS-CoV-2 RNA copies/ml), sensitivity was 96.6% (28/29; 95% CI 82.8-99.8) for both self- and professional testing. Deviations in sampling and testing (incomplete self-sampling or extraction procedure, or imprecise volume applied on the test device) were observed in 25 out of the 40 PCR-positives. Participants were rather young (mean age 35 years) and educated (59.6% with higher education degree). Most participants (80.9%) considered the Ag-RDT as rather easy to perform.

**Conclusions:** Ambulatory participants suspected for SARS-CoV-2 infection were able to reliably perform the Ag-RDT and test themselves. Procedural errors might be reduced by refinement of the Ag-RDTs for self-testing, such as modified instructions for use or product design/procedures. Self-testing may result in more wide-spread and more frequent testing. Paired with the appropriate information and education of the general public about the benefits and risks, self-testing may therefore have significant impact on the pandemic.

## INTRODUCTION

Antigen-detecting rapid diagnostic tests (Ag-RDTs) enable new COVID-19 risk mitigation strategies due to their short turn-around time and ease-of-use [1, 2]. Accordingly, resource-efficient screening and serial testing, e.g., in healthcare facilities, nursing homes, schools, or high-risk congregate settings, becomes feasible. However, Ag-RDTs have lower sensitivity and specificity than the gold standard reverse transcription-polymerase chain reaction (RT-PCR) [3]. Ag-RDTs have highest sensitivity in individuals with higher viral loads, typically observed during the first week of infection, the likely most-infectious period [4, 5]. Thus, Ag-RDTs may be suitable to detect most contagious cases, especially if performed frequently [6-8].

Most SARS-CoV-2 Ag-RDTs rely on nasopharyngeal (NP) sampling, requiring conduct by qualified healthcare workers and protective equipment. NP-sampling is frequently perceived as uncomfortable and, therefore, compliance with potential serial testing may be insufficient.

The term nasal sampling is often not used uniformly but can be differentiated in anterior nasal sampling (entire absorbent tip of the swab, usually 1 to 1.5 cm, inserted into nostril), and nasal mid-turbinate (swab inserted about 2 cm into nostril parallel to the palate until resistance is met at turbinates) [9]. Recent studies have shown the equivalence of nasal mid-turbinate (NMT) compared to NP-sampling for a WHO-listed SARS-CoV-2 Ag-RDT and established the feasibility of self-sampling under supervision [10, 11].

Self-testing, where a person collects his or her own specimen, performs the test and interprets the result, is an approach used for example in HIV control. There was initial concern that HIV self-tests may show reduced performance in the hands of lay persons, which proved to be unfounded. A systematic literature review concluded that self-testers can reliably and accurately perform HIV-RDTs [12].

For SARS-CoV-2, one Ag-RDT and one molecular RDT (based on loop mediated isothermal amplification) recently received an emergency use authorization in the USA for self-testing at home [13, 14]. Considering the ease-of-use of SARS-CoV-2 Ag-RDTs, they lend themselves to self-testing. Self-testing for SARS-CoV-2 may enable more wide-spread and more frequent testing. Paired with the appropriate information and education of the general public about the benefits and risks, self-testing may therefore have significant impact on the pandemic.

The objective of this study was to establish a head-to-head comparison of diagnostic accuracy, user-acceptability and feasibility of Ag-RDT self-testing with Ag-RDT professional testing and RT-PCR.

## METHODS

### Study design and participants

This was a manufacturer-independent prospective study of diagnostic accuracy, user acceptability and feasibility of an Ag-RDT when performed by patients themselves, with using a self-collected nasal mid-turbinate sample. For comparison, professional NP-sampling with testing on Ag-RDT was performed on the same participant as well as combined oropharyngeal (OP)/NP-sampling for RT-PCR as a reference. The study took place at the ambulatory SARS-CoV-2 testing facility of Charité - Universitätsmedizin Berlin, Germany, from 30 November to 11 December 2020. Participants eligible for inclusion were adults with high suspicion of SARS-CoV-2 infection, based on 1) reported contact with a confirmed case and any compatible symptom, or 2) fever or impaired taste or smell irrespective of exposure. Participants had to be able to understand the written instructions in German or English, defined as a minimum Common European Framework of Reference for Languages (CEFR) level of B2 (upper intermediate). Language proficiency was judged based on participants’ information and assessment by the study physicians. Participants were excluded if any of the swabs could not be collected. Participants were enrolled according to laboratory capacity in a consecutive series.

### Index test Ag-RDT

The Ag-RDT evaluated was the STANDARD Q COVID-19 Ag Test (SD Biosensor, Inc. Gyeonggi-do, Korea), which is also distributed by Roche [15]. While the test is commercially available as NP-sampling kit, the nasal-sampling kit is currently for research use only. Test kits were provided with differing flocked swabs, whereby the nasal swab used for self-sampling is less flexible with a larger sampling surface. There were differences in the manufacturer’s instructions for use of the two kits, with a more elaborate extraction process and a higher volume of extracted specimen used for testing nasal-samples.

### Study procedures

After providing written informed consent, the participants received a written and illustrated instruction for self-sampling and self-testing in German or English (Supporting Information S1 and S2), adapted from the manufacturer’s instructions for use. Participants performed the procedures in a separate room without time restrictions. A mirror for self-sampling and a timer were provided. The procedures were observed by a study physician, without answering questions or providing corrections.

The use of the test kit was evaluated with participant and observer questionnaires (Supporting Information S3 and S4). Deviations from the instructed technique were recorded. The participant was allowed a second attempt if desired. NMT self-sampling (both sides) was followed by professional NP-sampling (through one nostril) for Ag-RDTs and combined OP/NP-sampling (through the other nostril) for RT-PCR. The Ag-RDTs were performed directly after sampling at point-of-care by participants and trained study physicians.

All Ag-RDT results were interpreted by two study physicians, each blinded to the result of the other, also in addition to the participant’s interpretation of the self-test. The second reader was also blinded to the corresponding pairs (NMT/NP) of Ag-RDTs belonging to one individual. The visual read-out of the Ag-RDT test band was categorized as negative, weak positive, positive, or strong positive.

### Self-sampling

The written and illustrated instruction (Supporting Information S1) included the following steps: 1) “Cover your nose with a tissue and blow once”; 2) “Remove the swabs from the packaging by pulling on both flaps of the plastic film. Only touch the swab at the handle, not at the tip with the cotton"; 3) “Tilt your head back slightly (angle approx. 70 degrees)”; 4) “Insert the swab into a nostril with the cotton swab first. Slowly move the swab forward about 2 cm according to the angle (parallel to the palate = towards the throat, not upwards) until you feel resistance, do not apply pressure”; 5) “Slowly rotate the swab around for 15 seconds (minimum 4 rotations), rubbing all sides of the swab against the inside of the nose”; 6) “Repeat the procedure with the same swab in the other nostril”. The precise minimum number of 4 rotations with a time component of 15 seconds differs from the CDC guidance for NMT sampling, which recommends several rotations against the nasal wall [9].

### Self-testing

All materials necessary to perform one Ag-RDT were provided to the participant in a package. The written and illustrated instruction (Supporting Information S2) included the following steps: 1) “Open the extraction buffer tube and insert the swab with the cotton part first”; 2) “While squeezing the buffer tube in the lower part, stir the swab more than 10 times”; 3) “Remove the swab while squeezing the sides of the tube in the lower part to extract the liquid from the swab”; 4) “Press the nozzle cap tightly onto the tube”; 5) “Open the foil pouch of the test card at the tear line and put the test device on the table”; 6) “Place the test horizontally in front of you”; 7) “Hold the tube vertically over the marked round field (not the rectangle result window)”; 8) “Squeeze the tube for applying 4 drops of extracted specimen to the specimen well of the test device”. After 15 minutes, the participant was asked to state his/her interpretation of the test result (positive, negative, invalid, or don’t know).

### Reference standard RT-PCR

Sampling for RT-PCR testing was performed with a combined OP/NP swab (eSwab from Copan with 1 ml Amies medium) as *per* institutional recommendations for routine testing. The Roche cobas SARS-CoV-2 assay (Pleasanton, CA United States) or the SARS-CoV-2 E-gene assay from TIB Molbiol (Berlin, Germany) was performed for RT-PCR at the central laboratory. Viral RNA concentrations were calculated using assay-specific Ct-values, based on external calibrations curves [3, 16]. Staff performing the Ag-RDTs were blinded to RT-PCR results and *vice versa*.

### Additional data collection, user acceptability and feasibility

Participants were asked about comorbidities, symptoms, and their duration and severity. A paper-based questionnaire (Supporting Information S3) assessed prior testing experience (swab for COVID-19 taken in the past, use of RDTs, use of medical home-tests (e.g., for diabetes), laboratory work); education (highest school degree, vocational degree (e.g., apprenticeship, technical college), higher education degree); occupation; and language proficiency (CEFR). Occupations were classified according to the International Standard Classification of Occupations (ISCO-08). User acceptability and feasibility of self-sampling and self-testing were determined by participants on a scale from 1 (easy) to 5 (difficult). Furthermore, personal factors making the procedure more difficult (e.g., visual impairment) and suggestions for improvement were recorded. Data were collected and managed using Research Electronic Data Capture (REDCap) tools [17].

### Statistical analysis

Positive and negative percent agreements (PPA, NPA), sensitivity and specificity with 95% confidence intervals (95% CIs) were calculated for self and professional Ag-RDT use and compared against the reference standard RT-PCR [18]. The study was continued until 30 positive NP or NMT samples according to Ag-RDT were obtained, which is the minimum recommended by the WHO Emergency Use Listing Procedure to demonstrate sample type equivalency [19]. The targeted study size was 150 individuals (recent SARS-CoV-2 prevalence of 20% at testing site). Invalid Ag-RDT or RT-PCR results were reported separately. Descriptive statistics were used for participant characteristics, and user acceptability and feasibility assessment. All analyses were performed using R version 4.0.3.

### Ethics

This study was approved by the ethics committee of Charité - Universitätsmedizin (EA1/371/20).

## RESULTS

### Participants

The study flow is shown in Figure 1. Three participants were excluded from the study as they did not fulfil the CEFR minimum language criterion. After one further exclusion (lost PCR specimen), 146 participants were included in the analysis. Of these, 40 participants (27.4%) tested positive by RT-PCR.

**Fig. 1.**
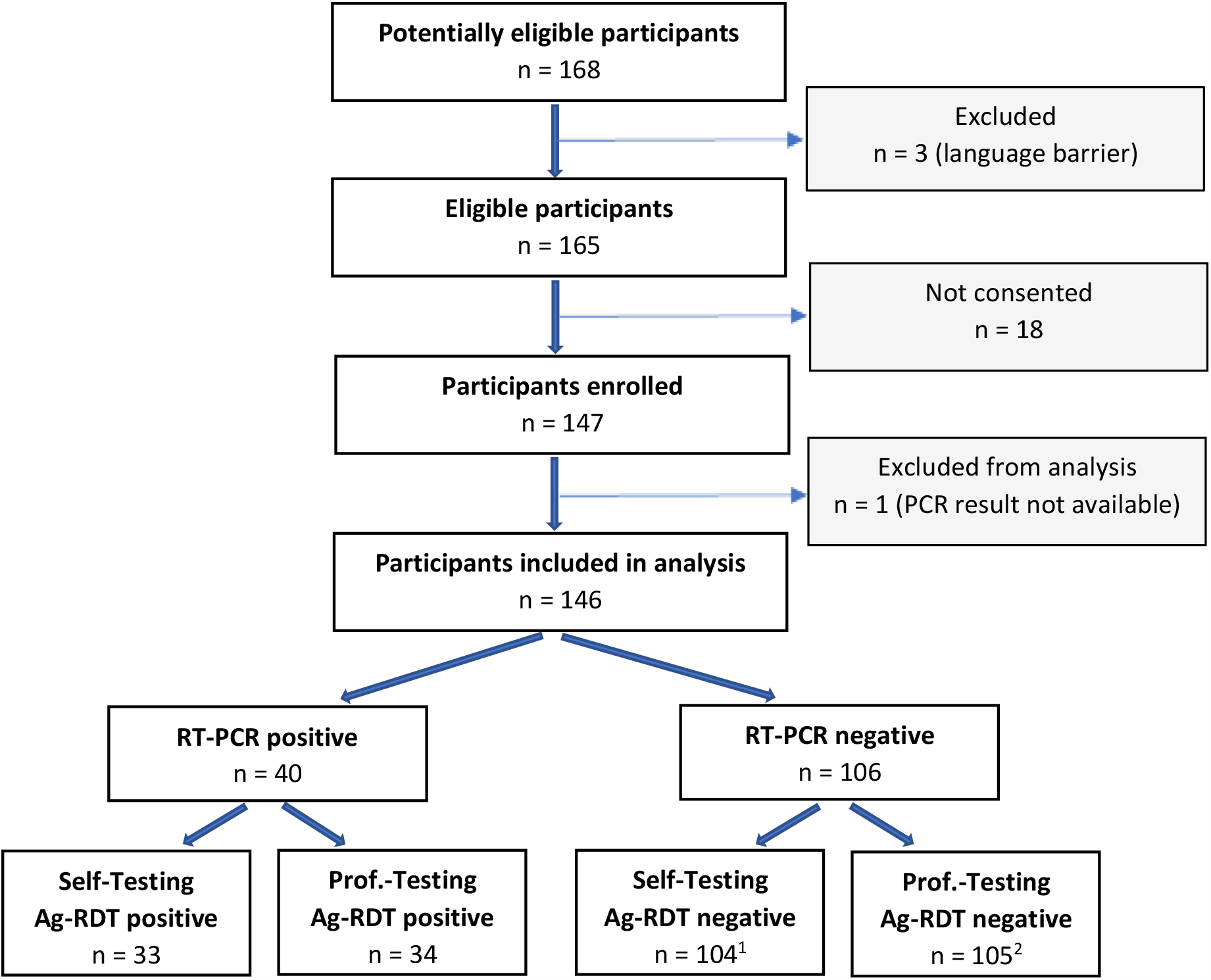
Study flow diagram. ^1^ one additional participant with invalid test result and one without test result (left before read-out) ^2^ one additional patient with false positive result

The characteristics of the participants are shown in Table 1. Mean age was 35 years (Standard Deviation [SD] 11.5), and 51.4% were female. All participants were symptomatic at the time of presentation, with a mean duration of 3.4 days (SD 2.0) post symptom onset. Sixty-six participants (46.2%) had a prior swab for SARS-CoV-2 testing collected in the past. Previous use of any RDTs and/or laboratory and/or home-test experience was reported by 29 participants (20.4%). An upper secondary school qualification was present in 108 (82.4%) participants, and 84 (59.6%) had a higher education degree. Twelve individuals (8.4%) were health or health associate professionals. Thirty-eight participants (26.6%) were not native German or English speakers. Among these, the German or English CEFR levels were B2 (upper intermediate) in 12, C1 (advanced) in 16, and C2 (mastery) in 10 individuals.

**Table 1.** Baseline demographics and characteristics of participants.

|  |  |  |
| --- | --- | --- |
| **Age in years** N=146 | Mean (SD) | 35.12 (11.5) |
| **Gender** N=146<br>- female<br>- male | n (%) | 75 (51.4%)<br>71 (48.6%) |
| **Comorbidities** N=146 | n (%) | 34 (23.3%) |
| **Symptoms on testing day** N=146 | n (%) | 146 (100%) |
| **Duration of symptoms in days** N=146 | Mean (SD) | 3.4 (2.0) |
| **Swab for COVID-19 taken in the past** N=143 | n (%) | 66 (46.2%) |
| **Previous RDT experience** N=141 | n (%) | 14 (9.8%)<sup>1</sup> |
| **Previous laboratory experience** N=142 | n (%) | 20 (14.1%)<sup>1</sup> |
| **Previous home-test experience** N=141 | n (%) | 4 (2.8%)<sup>1</sup> |
| **Highest general school qualification** N=131<br>- lower secondary school<br>- intermediate secondary school<br>- upper secondary school | n (%) | 0<br>23 (17.6%)<br>108 (82.4%) |
| **Highest further education** N=141<br>- none<br>- vocational degree<br>- higher education degree | n (%) | 22 (15.6%)<br>35 (24.8%)<br>84 (59.6%) |
| **Profession** N=143<br>- health professionals<br>- health associate professionals<br>- teaching professionals<br>- technicians and associate professionals<br>- other professionals<br>- clerical support, service, sales workers<br>- craft and related trades workers<br>- trainees, students<br>- others | n (%) | 9 (6.3%)<br>3 (2.1%)<br>27 (18.9%)<br>11 (7.7%)<br>25 (17.5%)<br>27 (18.9%)<br>7 (4.9%)<br>25 (17.5%)<br>9 (6.3%) |
| **Instruction in non-native language** N=143 | n (%) | 38 (26.6%) |
| **CEFR language level (English/German)** N=38<br>- B2 (upper intermediate)<br>- C1 (advanced)<br>- C2 (mastery) | n (%) | 12 (31.6%)<br>16 (42.1%)<br>10 (26.3%) |

<sup>1</sup> n=29 (20.4%) with previous RDT and/or laboratory and/or home-test experience

### Ag-RDT self- versus professional testing

Among 146 participants, 145 completed the testing (one left without reading the test-result). One patient did not want to repeat the test after spilling the buffer solution and obtaining an invalid result. No further invalid Ag-RDT results were recorded, and no participant required a second attempt. Results of self- and professional testing for 40 RT-PCR confirmed positive participants are shown in Table 2.

**Table 2.**
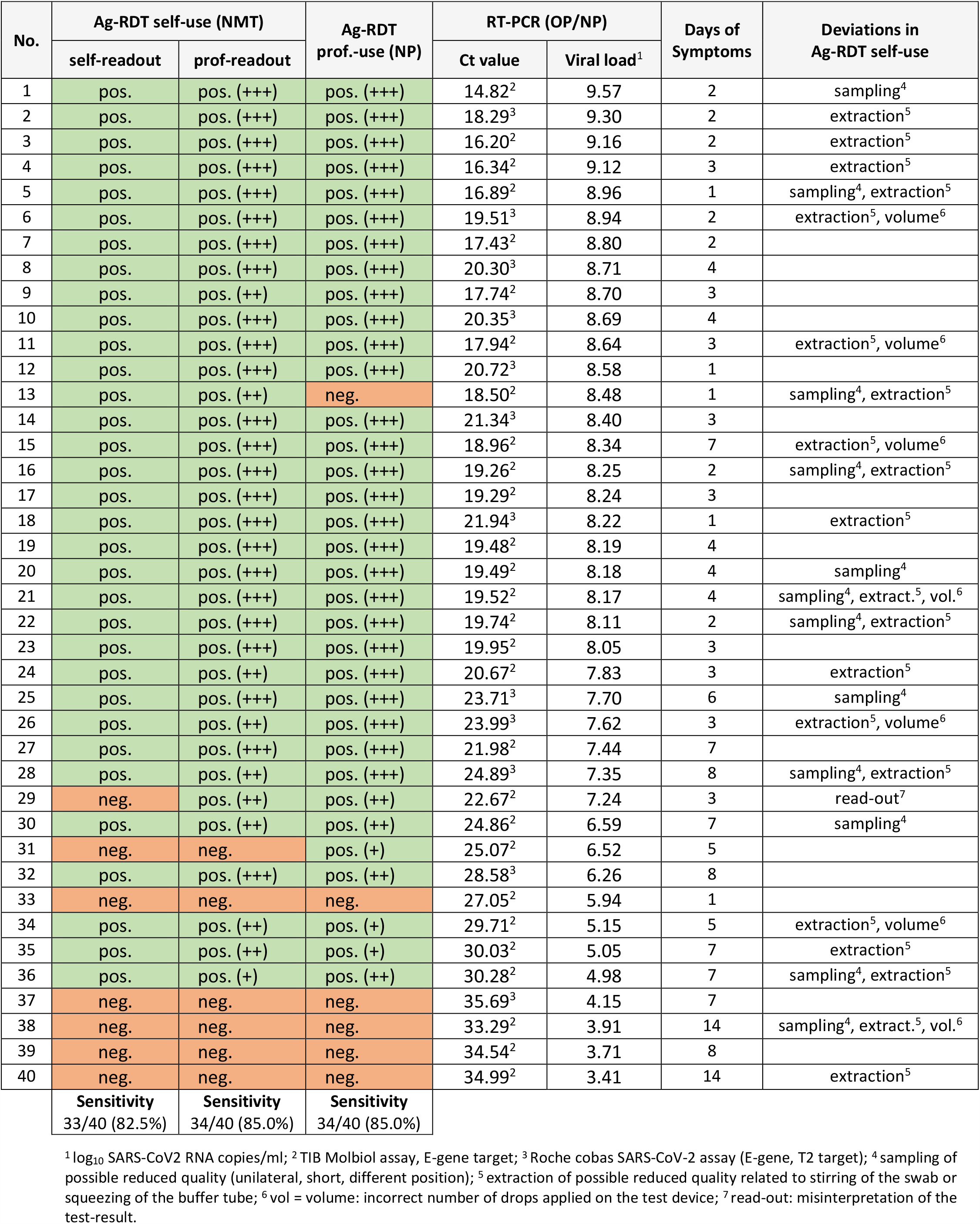
**Antigen-RDT self-testing (NMT sample) with self-readout and professional readout of the result, as well as professional testing (NP sample) in RT-PCR positive outpatients from combined OP/NP swab. Ct-values and viral loads of the paired RT-PCR samples are shown as well as the duration of symptoms. Relevant protocol deviations in the self-test are noted**.

Self-testing (including self-read-out) yielded a sensitivity of 82.5% (33/40 RT-PCR positives detected; 95% CI 68.1-91.3) and a specificity of 100% (95% CI 96.5-100) compared to RT-PCR. The sensitivity with professional Ag-RDT testing was 85.0% (34/40; 95% CI 70.9-92.9) and specificity was 99.1% (95% CI 94.8-99.5). In patients with high viral load (≥7.0 log_10_ SARS-CoV-2 RNA copies/ml) the sensitivity was 96.6% (28/29; 95% CI 82.8-99.8) for both self-testing and professional testing. In those with low viral load (<7.0 log_10_ SARS-CoV-2 RNA copies/ml) the sensitivity was 45.6% (5/11; 95% CI 21.3-72.0) for Ag-RDT self-use and 54.5% (6/11; 95% CI 28.0-78.7) for Ag-RDT professional use.

The positive percent agreement between self-testing (with self-read-out) and professional testing on Ag-RDT was 91.4% (32/35; 95% CI 77.6-97.0); the negative percent agreement was 99.1% (108/109; 95% CI 95.0-100). One patient (No. 31) was diagnosed by professional testing but not by self-testing. One patient (No. 29) with a positive self-test had falsely interpreted his result as negative. One patient (No. 13), who had a non-optimal NP swab due to poor tolerance, was diagnosed by self-testing only. One patient was false positive on Ag-RDT professional use.

Inter-rater reliability for the double read-out of the self-test by the participant and the study physician was high (kappa 0.98) and close to that of two study-physicians interpreting the self-test (kappa 1) or the professionally performed test (kappa 0.98). The semi-quantitative read-out after 15 minutes was more often higher for the professional NP testing versus the professional read-out of the self-testing (6 higher on professional testing, 4 higher on self-testing).

In a sub-analysis, the PPA of Ag-RDT self- and professional testing were similar for patients who had already undergone a swab for SARS-CoV-2 testing in the past (93.3%; 95% CI 70.0-99.7) and who did not (89.5%; CI 68.6-97.1).

### Feasibility of self-testing

Table 3 summarizes the individual steps of NMT self-sampling and self-testing with the proportion of participants who precisely followed the instructions. Deviations of self-sampling included a more vertically-directed angle for sampling (n=34), incorrect depth of insertion of the swab (n=6 too superficial, n=15 too deep), and reduced intensity of swabbing (as to duration n=35, rotations n=14 and rubbing n=61). Nine participants performed only unilateral NMT-sampling. One intervention by the study physician was necessary because of a possible risk of injury when the patient tried to insert the swab upside down into the nose.

**Table 3.** The relevant steps of self-sampling and self-testing with numbers of participants who precisely followed the instructions, according to observations of study physicians.

|  |  | Participants following precisely instructions |
| --- | --- | --- |
| **Self-sampling**<br>- removal of the swab from packing N=146<br>- swab only touched at handle N=146<br>- reclination of head N=146<br>- insertion about 2 cm N=146<br>- angle of the sampling direction N=145<br>- rotation for minimum 15 seconds N=145<br>- rotation minimum 4 times N=145<br>- rubbing against the nasal walls N=146<br>- bilateral sampling N=144 | n (%) | 125 (85.6%)<br>145 (99.3%)<br>129 (88.4%)<br>125 (85.6%)<br>121 (83.4%)<br>110 (75.9%)<br>131 (90.3%)<br>85 (58.2%)<br>135 (93.8%) |
| **Self-testing**<br>- opening of buffer tube N=142<br>- insertion of the swab into the tube N=145<br>- stirring more than 10 times N=146<br>- concurrent squeezing of the tube N=146<br>- swab removal while squeezing the tube N=145<br>- nozzle cap onto tube N=145<br>- tube held vertically N=144<br>- 4 drops applied N=145<br>- application to the specimen well N=144<br>- accurate interpretation N=145 | n (%) | 142 (100%)<br>145 (100%)<br>140 (95.9%)<br>95 (65.1%)<br>97 (66.9%)<br>144 (99.3%)<br>124 (86.1%)<br>99 (68.3%)<br>144 (100.0%)<br>144 (99.3%)<sup>1</sup> |

<sup>1</sup> one patient left before read-out

Deviations of self-testing were observed for the specimen extraction (less stirring of the swab n=6, inadequate tube squeezing while stirring n=51 and while removing the swab n=48). Furthermore, it proved difficult to apply exactly 4 drops to the sample well of the test device, with 40 participants applying more, and 6 less. Several drops coming out at once was the main problem.

Participants pointed out that their test performance may have been impaired by nervousness (n=5), fever or dizziness (n=2), poor concentration (n=2), feeling cold (n=2), aversion to self-sampling (n=2), language barrier (n=2) and limited fine motor skills (n=1).

On a scale from 1 (easy) to 5 (difficult), 114 (80.9%) participants stated that the test was rather easy to perform (scale 1/2); 23 (16.3%) found it medium easy/difficult (scale 3), and 4 (2.8%) rather difficult (scale 4).

The participants made the following suggestion to facilitate self-testing: instructions in a more precise or simple language (n=12), more illustrations (n=11), video format (n=9), instructions in additional languages (n=4), a mark on the swab to guide insertion depth (n=8), a less invasive sampling method (n=3, of which 2 preferred OP-sampling), information that sampling may feel uncomfortable (n=2), more general background information (n=2), and simplified unpacking of the swab (n=1).

## DISCUSSION

In this study among symptomatic outpatients, the sensitivities in detecting SARS-CoV-2 infection with an Ag-RDT were similar when patients performed NMT self-sampling and -testing (82.5%) as compared to professional NP-sampling and testing (85%). Self-testing and professional testing reached a high positive percent agreement (91.4%). At high viral load (>7.0 log_10_ SARS-CoV-2 RNA copies/ml), sensitivity was 96.6% for both approaches. Only one false-negative interpretation of a true positive self-test result occurred.

Sensitivities obtained in the present study were comparable to those observed in recent independent validation studies for the same Ag-RDT applied at this testing facility (73.2% to 80.5%) [10, 11]. The slightly higher sensitivities in this study are likely due to the higher number of patients with a high viral load (72.5% with ≥7.0 log_10_ SARS-CoV-2 RNA copies/ml, versus 48.8% and 64.1% in the prior studies). Differences in the test procedure of the nasal-sampling kit and NP-sampling kit, according to the instructions for use, may have influenced the sensitivities.

Notably, sensitivity was high despite frequent deviations from instructions (Tables 2 and 3). Regarding divergent Ag-RDT results between self-testing and professional testing, there was one participant yielding a false-negative result with self-testing (No. 31) who performed the test according to instructions, and one false-negative result with professional testing despite a high viral load (No. 13). In the latter patient, NP-sampling was not optimal due to poor tolerance, which occurs rather frequently in clinical practice. The recorded sensitivities are also remarkable considering the presumed limitations due to the acute illness and reported nervousness of patients. Furthermore, more than a quarter of the participants had a possible language barrier. This was reflected in the suggestions for improvement from the participants, which include the use of simple and clear language for instructions in additional languages, with more illustrations and a video format. Nevertheless, for most participants (80.9%) the Ag-RDT was rather easy to perform.

The strengths of the study are the rigorous methods, including standardized sampling, two independent blinded readers, an additional semi-quantitative assessment of Ag-RDT results, and the observation of procedures without intervention. The study is limited as it was performed in a single centre. Patients were rather young and educated, and almost half had experienced professional sample collection for SARS-CoV-2 in the past. In a sub-analysis, the PPA was similar regardless of whether a patient had previously experienced professional sample collection or not. The generalisability of the findings and applicability to settings with different prevailing patient characteristics needs to be confirmed. Further research should also be directed towards the interest and acceptability of using a self-test in different global regions.

The feasibility of self-testing is currently the subject of lively debate [8, 20, 21]. While in the US, the first home self-test was recently approved, but requires video-observation of the procedure, in Germany, Ag-RDTs for self-testing are available for limited populations including medical and care professionals, as well as for teachers and educators [6, 22]. Reasons to limit more widespread and unsupervised use of these tests include doubt as to the reliable conduct by lay people potentially giving rise to false-negative results, and false security resulting in unwitting transmission. False-positive results are considered a lesser problem, particularly when RT-PCR-confirmation is possible.

The data presented here demonstrates the feasibility and accuracy of self-testing in an unselected population and lay the ground for potential broader use. However, self-testing should be accompanied by widespread public campaigns informing about limited sensitivity, the importance of complementary hygiene measures, e.g., mask use, physical distancing, and the necessity of self-quarantine in case of a positive test. Reporting requirements in positive cases should be clarified. As long as Ag-RDTs test capacities are limited, deployment must be prioritised, also with a global perspective. Individually, Ag-RDT self-testing could contribute to resumption of a certain degree of normal life, e.g., self-testing could be done before visiting nursing homes. For society at large, recent modelling data suggest that repeated screening for SARS-CoV-2, combined with immediate reporting, isolation, and quarantine, can greatly reduce viral transmission. In this model, test sensitivity is of minor importance [7]. Self-testing with Ag-RDTs could not only alleviate overstretched RT-PCR capacity and medical personnel, but also may result in increased access to frequent testing and significant impact on the pandemic [7, 23].

In conclusion, we demonstrate that symptomatic persons can reliably perform a SARS-CoV-2 lateral-flow Ag-RDT and test themselves. Procedural errors might be reduced by refinement of the Ag-RDTs for self-testing, such as modified instructions for use or product design/procedures.

## Data Availability

All raw data and analysis code are available upon request to the corresponding author.

## Supporting Information

**S1** Instruction for self-sampling.

**S2** Instruction for self-testing.

**S3** Participant questionnaire.

**S4** Observer questionnaire.

## Author contributions

AKL, ON, and CMD designed the study and developed standard operating procedures. AKL, ON, and CR implemented the study design. ON, CR, MH, enrolled participants, performed laboratory work. AKL, CMD, and FPM led the writing of the manuscript. FPM and JS coordinated and supervised the study site. MW, FK, JB, and SB enrolled participants. MGe coordinated the testing facility. FT and MGa led the data analysis. FL provided technical advice. VMC and TCJ were responsible for PCR testing and contributed to the interpretation of the data. JAS supported the study design setup. All authors have reviewed the manuscript.

## Acknowledgements

Heike Rössig, Elisabeth Linzbach, Katja von dem Busche, Stephanie Padberg, Melanie Bothmann, Zümrüt Tuncer, Stefanie Lunow, Beate Zimmer, Astrid Barrera Pesek, Sabrina Pein, Verena Haack, Oliver Deckwart, Birgit Zittlau, Andrea Junge.

## Data availability

All raw data and analysis code are available upon request to the corresponding author.

