## Supporting Information S1 for "SARS-CoV-2 patient self-testing with an antigen-detecting rapid test: a head-to-head comparison with professional testing"

UniversitätsKlinikum Heidelberg

CHARITÉ

### Instruction
### nasal self-swab

(Version vom 30.11.2020)

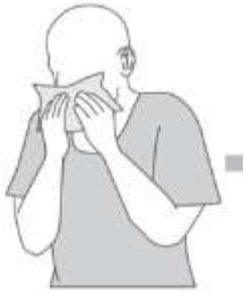

1. Cover your nose with a tissue and blow once

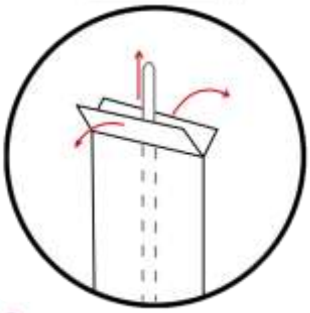

2. Remove the swabs from the packaging by pulling on both flaps of the plastic film. Only touch the swab at the handle, not at the tip with the "cotton swab".

3. Tilt your head back slightly (angle approx. 70 degree).

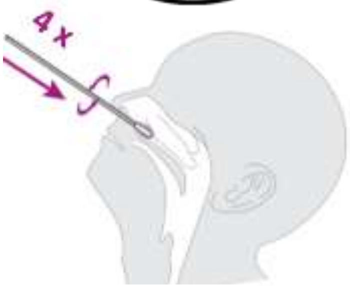


4. Insert the swab into a nostril with the "cotton swab" first. Slowly move the swab forward about 2 cm according to the angle (parallel to the palate = towards the throat, not upwards) until you feel resistance, do not apply pressure.

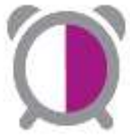


5. Slowly rotate the swab around for 15 seconds (minimum 4 rotations), rubbing all sides of the swab against the inside of the nose.

6. Slowly remove the swab.

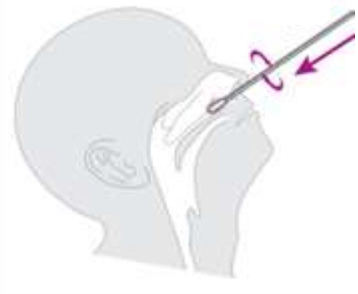

7. Repeat the procedure with the same swab in the other nostril.

8. Place the swab in the dish.

**We would like to observe how you take the smear yourself**
**- without answering questions**
