## Supporting Information S2 for "SARS-CoV-2 patient self-testing with an antigen-detecting rapid test: a head-to-head comparison with professional testing"

### Instruction Self-Test-Study

Version 1 – 30.11.20

English

### Please check if the following test components are available

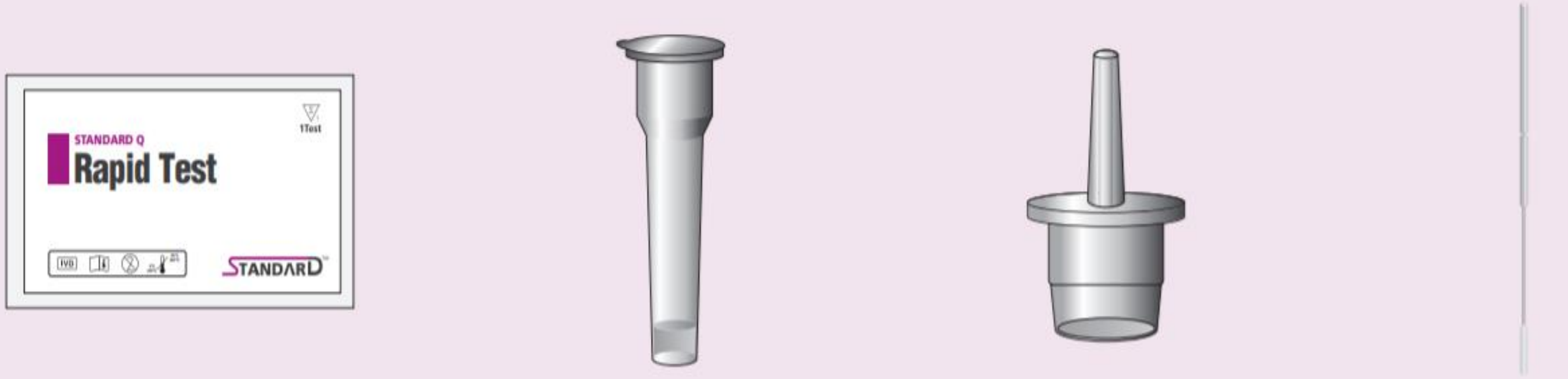


**Test device**
(individually in a foil pouch with desiccant)

**Extraction buffer tube**

**Nozzle cap**

**Sterile swab**

### Test procedure I

1. Open the extraction buffer tube and insert the swab **with the cotton part first.**

2. While **squeezing the buffer tube in the lower part**, **stir the swab more than 10 times**. (1).

3. **Remove** the swab **while squeezing the sides of the tube in the lower part** to extract the liquid from the swab (2).

4. Press the nozzle cap tightly onto the tube (3).

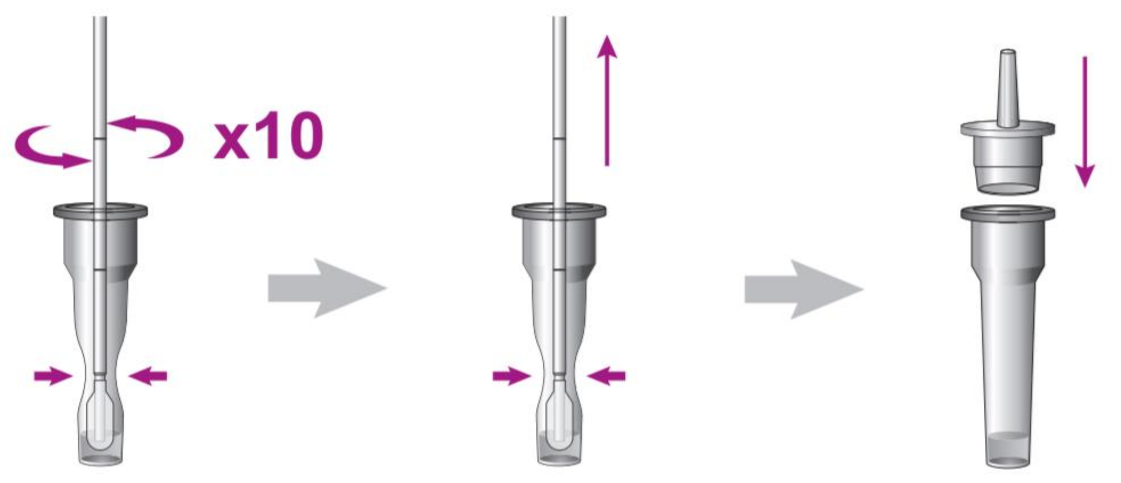


(1) (2) (3)

### Test procedure II

5. Check the expiry date on the back of the foil pouch (1).

6. Open the foil pouch of the test card at the **tear line** (2) and put the test device on the table.

7. Ensure that the **desiccant** status indicator shows valid (yellow) (4).

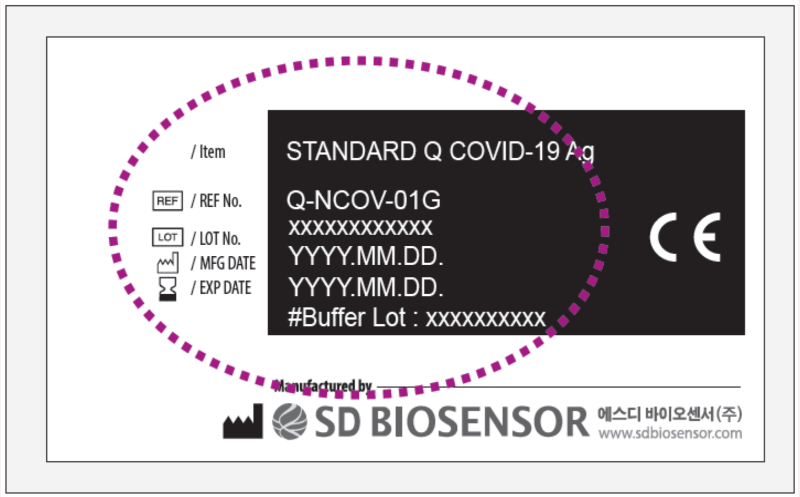


(1)

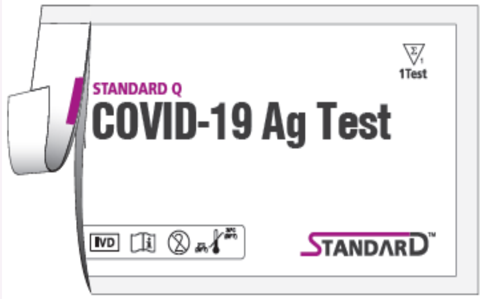


(2)

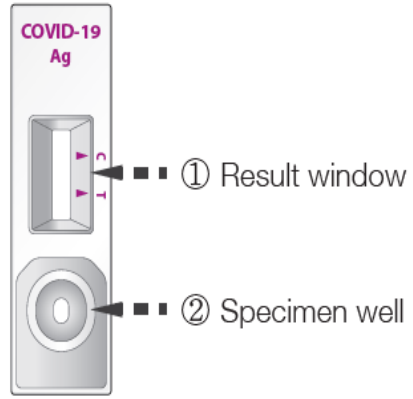


(3)

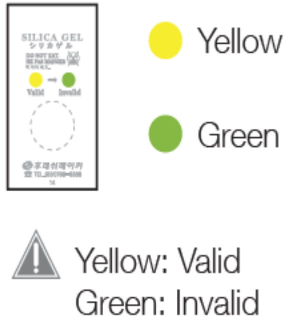


(4)

### Test procedure III



8. Place the test **horizontally** in front of you as shown in figure (1).

9. Hold the tube **vertically over the marked round field** (<u>**not**</u> the rectangle result window) (1).

10. Squeeze the tube for applying **4 drops** of extracted specimen to the specimen well of the test device (1). (You can also continue the test if you have accidentally applied 5 drops)

11. Set the timer and read test result at 15 min (2).

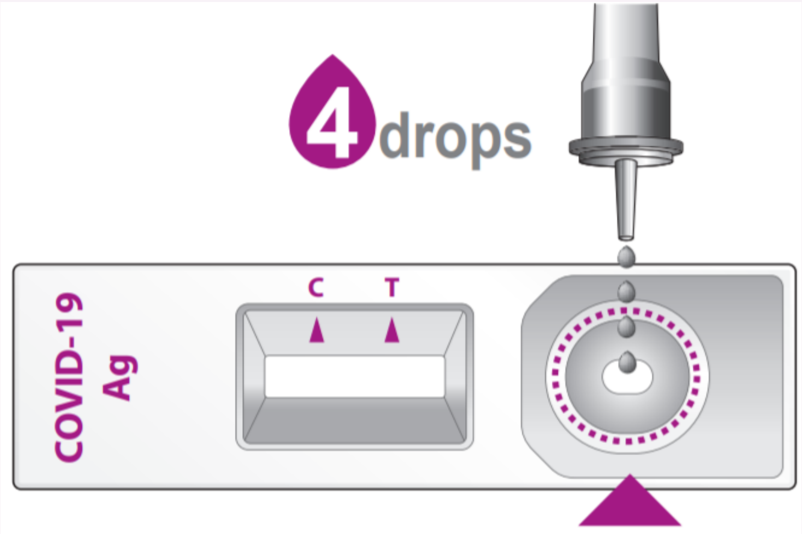


(1)

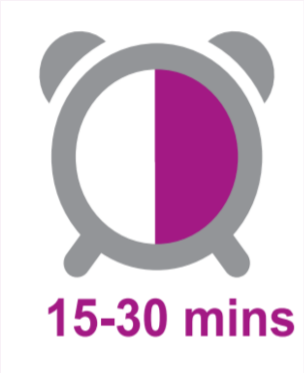


(2)

### Interpretation

| Test Result | Example | Description |
|---|---|---|
| **Negative** | 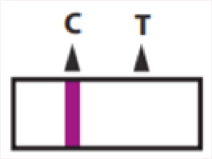  | **Negative:** A colored band will appear in the left section of the result window to show that the test is working properly. This band is control line (C).<br>-> no sign of infection |
| **Positive** | 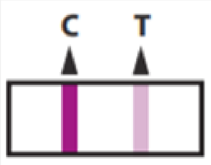  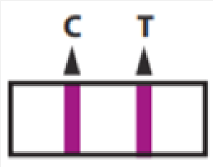  | **Positive:** A colored band will appear in the right section of the result window. This band is test line of SARS-CoV-2 antigen (T).<br>-> sign of infection |
| **Invalid** | 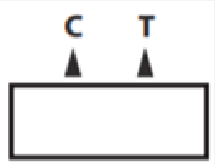  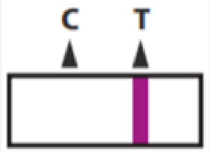  | **Invalid:** If no control line (C) is visible, the result is considered invalid.<br>-> the test is not working properly |

**<u>-> Also pay attention to faint lines <-</u>**

Weak (pale) or irregular lines should also be considered.
