## Supporting Information S3 for "SARS-CoV-2 patient self-testing with an antigen-detecting rapid test: a head-to-head comparison with professional testing"

Studien-ID

### Questionnaire self-test COVID-19

(Version vom 06.12.2020)

Dear participant,

We are very thankful for your participation in the study. While waiting for the result of the antigen test ("rapid test"), please answer the following questions. We will evaluate your data pseudonymously, without being able to draw any conclusions about your person. **Thank you!**

#### 1. Experiences

- Profession: ____________________________

- Did you have a COVID-19 test with a swab before?

  ❑YES ❑NO

- Do you have experience in conducting rapid tests?

  ❑YES ❑NO

- Do you have any laboratory working experience?

  ❑YES ❑NO

- Do you regularly use a medical test at home (e.g. blood sugar measurement, blood coagulation measurement, HIV self-test)?

  ❑YES ❑NO

#### 2. Test evaluation *scale from 1 to 5, only one mark per question*

- How easy/difficult did you find it **to perform** the rapid antigen test?

  ❑1 (1=**easy**) ❑2 ❑3 ❑4 ❑5 (5=**difficult**)

- How understandable did you find the **instructions**?

  ❑1 (1=**easy**) ❑2 ❑3 ❑4 ❑5 (5=**difficult**)

- How easy/difficult do you think **most people** can **learn to use** the rapid antigen test?

  ❑1 (1=**easy**) ❑2 ❑3 ❑4 ❑5 (5=**difficult**)

- Was there a **personal reason** that made it **difficult** for you to perform the test? (e.g. visual impairment, disturbance of fine motor skills)?

  ______________________________________________________________________

- Do you have a **suggestion for improvement** to simplify the test?

  ______________________________________________________________________

#### 3. School / education

- What is your highest **general school qualification**?

  ❑ lower secondary school (Hauptschule)

  ❑ intermediate secondary school (mittlerer Schulabschluss)

  ❑ (specialised) upper secondary school (Fachhochschulreife, Abitur)

  ❑ other: ______________________________

- Do you have a **certificate of education for a certain profession / vocational degree**? [e.g. apprenticeship (Lehre), vocational school (Berufsfachschule), technical college, training for civil servants)?

  ❑YES, namely ______________________________ ❑NO

- Do you have a **higher education degree**? [e.g. university, specialised college of higher education (Fachhochschule) college of advanced vocational studies (Berufsakademie)]

  ❑YES, namely ______________________________ ❑NO

- **Other** educational degree: ______________________________________________

#### 4. Language

- What is your **native language**?

  ❑ English ❑ other: ______________________________

- If you have another mother tongue, how good is your use of English?

  ❑ **Basic** user
  - ❑ A1: beginner
  - ❑ A2: elementary

  ❑ **Independent** user
  - ❑ B1: intermediate and
  - ❑ B2: upper intermediate

  ❑ **Proficient** User
  - ❑ C1: advanced
  - ❑ C2: mastery
