## Supporting Information S4 for "SARS-CoV-2 patient self-testing with an antigen-detecting rapid test: a head-to-head comparison with professional testing"

**Institut für Tropenmedizin und Internationale Gesundheit**

Komm. Direktor: Herr Prof. Dr. med. Frank Mockenhaupt

http://tropeninstitut.charite.de/

### Self-Test-Study: Observation form

Observer: ______________ Study-ID:______________ Date: ______________

#### Part A – Selfswab of the anterior nose

- ✓ Important: No verbal or non-verbal assistance/correction should be given
- ✓ Exception: Actively encourage hand disinfection

| Steps | Observation | | Comment |
|---|---|---|---|
| 1. Swab opened correctly (flaps)? | ❑YES | ❑NO | |
| 2. Touching the tip? | ❑YES | ❑NO | |
| 3. Head reclination approx. 70 degree? | ❑YES | ❑NO | |
| 4. Depth approx. 2 cm? | ❑YES | ❑NO | |
| 5. Correct angle/direction? | ❑YES | ❑NO | |
| 6. At least 15 sec rotation? | ❑YES | ❑NO | |
| 7. At least 4x rotation? | ❑YES | ❑NO | |
| 8. Rubbed against nose walls? | ❑YES | ❑NO | |
| 9. Both sides? | ❑YES | ❑NO | |
| **Other:** | | | |

**Institut für Tropenmedizin und Internationale Gesundheit**

Komm. Direktor: Herr Prof. Dr. med. Frank Mockenhaupt

http://tropeninstitut.charite.de/

#### Part B – Performing an Ag-RDT

- ✓ Important: No verbal or non-verbal assistance/correction should be given
- ✓ 1. Exception: Actively encourage hand disinfection

  2. Test result is requested from the study participant at the end

| Steps | Observation | | Comment |
|---|---|---|---|
| 1. Checked test components? | ❑YES | ❑NO | |
| 2. Opened flap of extraction buffer tube? | ❑YES | ❑NO | |
| 3. Inserted swab correctly in buffer tube? | ❑YES | ❑NO | |
| 4. Squeezed the lower part of the buffer tube? | ❑YES | ❑NO | |
| 5. Rotated the swab at least 10 times meanwhile? | ❑YES | ❑NO | |
| 6. Squeezed the cotton part before removing? | ❑YES | ❑NO | |
| 7. Pressed nozzle cap tightly onto tube? | ❑YES | ❑NO | |
| 8. Checked expiry date? | ❑YES | ❑NO | |
| 9. Opened foil pouch on the tear line? | ❑YES | ❑NO | |
| 10. Checked desiccant? | ❑YES | ❑NO | |
| 11. Held tube vertically during application? | ❑YES | ❑NO | |
| 12. Correct amount of drops (4)? | ❑YES | ❑NO | |
| 13. – dropped into correct field (specimen well, not result window)? | ❑YES | ❑NO | |
| 14. Set the timer? | ❑YES | ❑NO | |
| 15. Read test result after 15 min? | ❑YES | ❑NO | |
| 16. Interpretated test result correctly? | ❑YES | ❑NO | |

CHARITÉ

**Institut für Tropenmedizin und Internationale Gesundheit**

Komm. Direktor: Herr Prof. Dr. med. Frank Mockenhaupt

http://tropeninstitut.charite.de/

| | | |
|---|---|---|
| 17. Second try? If yes, why? | ❑YES | ❑NO |
| 18. Other deviations? If yes, which? | ❑YES | ❑NO |
| 19. Linguistic comprehension problems of the instructions? | ❑YES | ❑NO |
| 20. Content-related comprehension problems of the instructions? | ❑YES | ❑NO |
| 21. Language level **German** ❑ or **English** ❑ (if not native speaker)? | ❑ A1 ❑ A2<br>❑ B1 ❑ B2<br>❑ C1 ❑ C2 | |

| | | |
|---|---|---|
| **Test result** | | How does the **study participant** interprets the test result ?<br>❑negative ❑positive ❑invalid ❑does not know<br>comment: |
| | | How does the **study physician** interprets the test result ?<br>❑negative ❑positive ❑invalid ❑does not know<br>comment: |
| **Application** | | How do you rate the level of **self-confidence** during the test performance?<br>❑1 (very uncertain) ❑2 ❑3 ❑4 ❑5 (very confident)<br>comment: |
| | | Do you consider the study person to be capable of **independent home testing** ?<br>☐yes ☐no ☐questionable ☐other: |
